# First 12 patients with coronavirus disease 2019 (COVID-19) in the United States

**DOI:** 10.1101/2020.03.09.20032896

**Authors:** The COVID-19 Investigation Team, Stephanie A. Kujawski, Karen K Wong, Jennifer P. Collins, Lauren Epstein, Marie E. Killerby, Claire M. Midgley, Glen R. Abedi, N. Seema Ahmed, Olivia Almendares, Francisco N. Alvarez, Kayla N. Anderson, Sharon Balter, Vaughn Barry, Karri Bartlett, Karlyn Beer, Michael A. Ben-Aderet, Isaac Benowitz, Holly Biggs, Alison M. Binder, Stephanie R. Black, Brandon Bonin, Catherine M. Brown, Hollianne Bruce, Jonathan Bryant-Genevier, Alicia Budd, Diane Buell, Rachel Bystritsky, Jordan Cates, E. Matt Charles, Kevin Chatham-Stephens, Nora Chea, Howard Chiou, Demian Christiansen, Victoria Chu, Sara Cody, Max Cohen, Erin Conners, Aaron Curns, Vishal Dasari, Patrick Dawson, Traci DeSalvo, George Diaz, Matthew Donahue, Suzanne Donovan, Lindsey M. Duca, Keith Erickson, Mathew D. Esona, Suzanne Evans, Jeremy Falk, Leora R. Feldstein, Martin Fenstersheib, Marc Fischer, Rebecca Fisher, Chelsea Foo, Marielle J. Fricchione, Oren Friedman, Alicia M. Fry, Romeo R. Galang, Melissa M. Garcia, Susa I. Gerber, Graham Gerrard, Isaac Ghinai, Prabhu Gounder, Jonathan Grein, Cheri Grigg, Jeffrey D. Gunzenhauser, Gary I. Gutkin, Meredith Haddix, Aron J. Hall, George Han, Jennifer Harcourt, Kathleen Harriman, Thomas Haupt, Amber Haynes, Michelle Holshue, Cora Hoover, Jennifer C. Hunter, Max W. Jacobs, Claire Jarashow, Michael A. Jhung, Kiran Joshi, Talar Kamali, Shifaq Kamili, Lindsay Kim, Moon Kim, Jan King, Hannah L. Kirking, Amanda Kita-Yarbro, Rachel Klos, Miwako Kobayashi, Anna Kocharian, Kenneth K. Komatsu, Ram Koppaka, Jennifer E. Layden, Yan Li, Scott Lindquist, Stephen Lindstrom, Ruth Link-Gelles, Joana Lively, Michelle Livingston, Kelly Lo, Jennifer Lo, Xiaoyan Lu, Brian Lynch, Larry Madoff, Lakshmi Malapati, Gregory Marks, Mariel Marlow, Glenn E. Mathisen, Nancy McClung, Olivia McGovern, Tristan D. McPherson, Mitali Mehta, Audrey Meier, Lynn Mello, Sung-sil Moon, Margie Morgan, Ruth N. Moro, Janna' Murray, Rekha Murthy, Shannon Novosad, Sara E. Oliver, Jennifer O'Shea, Massimo Pacilli, Clinton R. Paden, Mark A. Pallansch, Manisha Patel, Sajan Patel, Isabel Pedraza, Satish K. Pillai, Talia Pindyck, Ian Pray, Krista Queen, Nichole Quick, Heather Reese, Brian Rha, Heather Rhodes, Susan Robinson, Philip Robinson, Melissa Rolfes, Janell Routh, Rachel Rubin, Sarah L. Rudman, Senthilkumar K. Sakthivel, Sarah Scott, Christopher Shepherd, Varun Shetty, Ethan A. Smith, Shanon Smith, Bryan Stierman, William Stoecker, Rebecca Sunenshine, Regina Sy-Santos, Azaibi Tamin, Ying Tao, Dawn Terashita, Natalie J. Thornburg, Suxiang Tong, Elizabeth Traub, Ahmet Tural, Anna Uehara, Timothy M. Uyeki, Grace Vahey, Jennifer R. Verani, Elsa Villarino, Megan Wallace, Lijuan Wang, John T. Watson, Matthew Westercamp, Brett Whitaker, Sarah Wilkerson, Rebecca C. Woodruff, Jonathan M. Wortham, Tiffany Wu, Amy Xie, Anna Yousaf, Matthew Zahn, Jing Zhang

## Abstract

**Introduction:** More than 93,000 cases of coronavirus disease (COVID-19) have been reported worldwide. We describe the epidemiology, clinical course, and virologic characteristics of the first 12 U.S. patients with COVID-19.

**Methods:** We collected demographic, exposure, and clinical information from 12 patients confirmed by CDC during January 20–February 5, 2020 to have COVID-19. Respiratory, stool, serum, and urine specimens were submitted for SARS-CoV-2 rRT-PCR testing, virus culture, and whole genome sequencing.

**Results:** Among the 12 patients, median age was 53 years (range: 21–68); 8 were male, 10 had traveled to China, and two were contacts of patients in this series. Commonly reported signs and symptoms at illness onset were fever (n=7) and cough (n=8). Seven patients were hospitalized with radiographic evidence of pneumonia and demonstrated clinical or laboratory signs of worsening during the second week of illness. Three were treated with the investigational antiviral remdesivir. All patients had SARS-CoV-2 RNA detected in respiratory specimens, typically for 2–3 weeks after illness onset, with lowest rRT-PCR Ct values often detected in the first week. SARS-CoV-2 RNA was detected after reported symptom resolution in seven patients. SARS-CoV-2 was cultured from respiratory specimens, and SARS-CoV-2 RNA was detected in stool from 7/10 patients.

**Conclusions:** In 12 patients with mild to moderately severe illness, SARS-CoV-2 RNA and viable virus were detected early, and prolonged RNA detection suggests the window for diagnosis is long. Hospitalized patients showed signs of worsening in the second week after illness onset.

## INTRODUCTION

In December 2019, an outbreak of the novel disease COVID-19 caused by the newly identified severe acute respiratory syndrome coronavirus 2 (SARS-CoV-2) was reported in Wuhan City, Hubei province, China. As of March 4, 2020, more than 93,000 COVID-19 cases have been reported in 73 countries,^1^ and 80 confirmed and presumptive patients with COVID-19 have been identified in the United States;^2^ one has been previously described in detail.^3^

SARS-CoV-2 RNA detection, virus culture, and the relationship to the clinical course of COVID-19 are not fully understood. We report the epidemiology, clinical course, clinical management, and virologic characteristics of the first 12 patients with COVID-19 diagnosed in the United States.

## METHODS

### Case identification and confirmation

Local health departments in consultation with clinicians identified patients under investigation (PUI) for COVID-19 beginning January 17, 2020. PUI testing criteria changed during this period but included the presence of fever and/or lower respiratory symptoms (e.g., cough or shortness of breath) and at least one epidemiologic risk factor in the two weeks before symptom onset. During January 17–31, epidemiologic risk factors were history of travel from Wuhan City, close contact with an ill PUI, or close contact with a patient with laboratory-confirmed COVID-19.^4,5^ Beginning February 1, epidemiologic risk factors changed to close contact with a patient with confirmed COVID-19 or history of travel from mainland China.^6^ During both time periods, close contact was defined as being within 6 feet for a prolonged period of time^7^ or contact with respiratory secretions.^8^ Specimens from PUIs were tested for SARS-CoV-2 at the Centers for Disease Control and Prevention (CDC).

Upper respiratory tract specimens (nasopharyngeal [NP], oropharyngeal [OP]) and available lower respiratory tract specimens (sputum) were collected and tested for SARS-CoV-2 RNA by real-time reverse-transcription polymerase chain reaction (rRT-PCR).^3^ A case of COVID-19 was defined as identification of laboratory-confirmed SARS-CoV-2 in ≥ 1 specimen from a patient. We included patients with COVID-19 who were confirmed by CDC during January 20–February 5, 2020.

### Data and follow-up specimen collection

Patients with COVID-19 were interviewed by public health officials to collect information on demographics, exposures, travel history, and symptoms, including signs or symptoms before presentation. For all twelve patients, available medical records were reviewed. For hospitalized patients, clinicians systematically abstracted clinical data from the medical record.

Illness day 1 was defined as the first day of reported COVID-19 signs and symptoms; collection date of the first SARS-CoV-2-positive specimen was used for one patient with no clear symptom onset date. When symptoms at onset or onset dates in the medical record differed from those reported from the public health interview, the latter were used. Results for virologic tests were reported relative to illness day 1. Duration of potential exposure to SARS-CoV-2 was defined as dates of travel to China or dates of first to last exposure to a U.S. patient with COVID-19. Fever was defined as subjective fever or temperature ≥ 100.4 °F.

We requested collection of NP swabs, OP swabs, sputum (if available), serum, urine, and stool from each patient initially for every 2–3 days for the first 17 days of illness for SARS-CoV-2 virologic testing.^9^

Further specimens were collected for testing if the patient continued to test positive for SARS-CoV-2 beyond day 17.

CDC’s Human Research Protection Office determined this work was exempt from human subjects’ research regulations as it involved identification, control, or prevention of disease in response to an immediate public health threat. Forms used in this response were approved under OMB, number 0920-1011. Data were analyzed and visualized using Excel, SAS 9.4, R 3.6.2, and Python 3.7.3.^10–13^

### Laboratory Methods

Specimens were evaluated using SARS-CoV-2 RNA detection, virus culture, whole genome sequencing, and phylogenetic analysis. Virus culture was attempted from early SARS-CoV-2-positive respiratory specimens (NP swabs, OP swabs, and sputum) from 9 patients. Further virus culture is ongoing. Detailed methods are included in the Appendix.

## RESULTS

### Epidemiologic and clinical characteristics

Twelve patients with confirmed COVID-19 were identified in six states. Five patients received only outpatient care and were isolated at home (Patients 1–5), and seven were hospitalized (Patients 6–12) (Figure 1). Median patient age was 53 years (range: 21–68); eight patients were male (Table 1). Four of five patients with ≥ 1 underlying medical conditions were hospitalized (Tables 1 and 2).

**Table 1:** Demographic characteristics, exposure history, and clinical characteristics of the first 12 patients with COVID-19 in the United States, January–February 2020

|  | Values (N=12) |  |
| --- | --- | --- |
| Median age (range) – years | 53 | (21–68) |
|  | n/N | % |
| Male sex – no. | 8/12 | 67 |
| State – no. |  |  |
| California | 6/12 | 50 |
| Illinois | 2/12 | 17 |
| Arizona | 1/12 | 8 |
| Massachusetts | 1/12 | 8 |
| Washington | 1/12 | 8 |
| Wisconsin | 1/12 | 8 |
| Initial source of case identification – no. |  |  |
| Outpatient clinic/urgent care | 4/12 | 33 |
| Emergency department | 4/12 | 33 |
| Health department<sup>a</sup> | 2/12 | 17 |
| Port of entry | 1/12 | 8 |
| Close contact active monitoring | 1/12 | 8 |
| Exposure history (≤14 days before illness onset) |  |  |
| Travel to mainland China – no. | 10/12 | 83 |
| Travel to Wuhan, Hubei Province, China – no. | 9/12 | 75 |
| Exposure to a confirmed U.S. COVID-19 patient – no. | 2/12 | 17 |
| Underlying medical conditions – no. | 5/12 | 50 |
| Cardiac disease<sup>b</sup> – no. | 2/12 | 17 |
| Hypertension – no. | 2/12 | 17 |
| Diabetes mellitus – no. | 1/12 | 8 |
| Chronic lung disease – no. | 1/12 | 8 |
| High cholesterol – no. | 2/12 | 17 |
| Fatty liver disease – no. | 1/12 | 8 |
| Hepatitis B – no. | 1/12 | 8 |
| Current tobacco use – no. | 1/12 | 8 |
| Symptoms reported at illness onset<sup>c</sup> – no. |  |  |
| Fever | 7/12 | 58 |
| Subjective | 5/7 | 57 |
| Measured (≥100.4°F or 38°C) | 2/7 | 43 |
| Cough<sup>d</sup> | 8/12 | 67 |
| Fatigue | 5/12 | 42 |
| Shortness of breath (dyspnea) | 1/12 | 17 |
| Sore throat | 1/12 | 8 |
| Headache | 3/12 | 25 |
| Runny nose (rhinorrhea)/nasal congestion | 1/12 | 8 |
| Chills | 1/12 | 8 |
| Diarrhea | 1/12 | 8 |
| Nausea | 1/12 | 8 |
| Days of first sample collection, median (range) (n=12) | 4 | (1–9) |
| Highest level of healthcare utilization – no. |  |  |
| Outpatient clinic or urgent care | 2/12 | 17 |
| Emergency department | 3/12 | 25 |
| Hospitalized | 7/12 | 58 |
| Abnormal chest radiograph – no. | 7/9 | 78 |

<sup>a</sup> Includes one case who was a contact of a confirmed COVID-19 case
<sup>b</sup> Includes coronary artery disease, and pacemaker for bradycardia. Reports of hypertension were not included.
<sup>c</sup> Initial symptoms were obtained through patient interview, and may have differed from symptoms at the time of identification
<sup>d</sup> One patient reported a cough with initial onset in mid-December before the patient traveled to China. The patient reported no change in the cough from the initial onset until resolution 2 weeks after SARS-CoV-2 was first detected.

**Table 2:** Clinical characteristics of the first seven patients hospitalized with COVID-19 in the United States, January–February 2020*

| Characteristic | Patient 6 | Patient 7 | Patient 8 | Patient 9 | Patient 10 | Patient 11 | Patient 12 |
| --- | --- | --- | --- | --- | --- | --- | --- |
| Age group (years) | 30–39 | 60–69 | 60–69 | 30–39 | 50–59 | 50–59 | 50–59 |
| Sex | Male | Female | Male | Male | Male | Male | Female |
| Underlying medical conditions | Hypertriglyceridemia | Hypertension, hyperlipidemia, pacemaker for bradycardia | Tobacco use, hypertension, coronary artery disease, COPD, history of lung cancer status post partial lobectomy | Type 2 Diabetes mellitus, fatty liver | None | None | None |
| Reported symptoms on illness day 1 | Cough, subjective fever | Fatigue, subjective fever | Productive cough, worsening shortness of breath, fatigue, subjective fever | Diarrhea** | Dizziness, cough, nasal congestion, subjective fever | Fever, fatigue, cough | Fever, headache |
| rRT-PCR positive extrapulmonary sites | None | Stool | None | Serum, stool | None | Stool | Stool |
| Initial normal CXR (day) | Yes (4, 7) | Yes (7) | No | Yes (4) | Yes (9) | No | No |
| First abnormal chest imaging findings (day) | Left basilar opacity (9) | Multifocal infiltrates, mediastinal and hilar lymphadenopathy*** (7) | Right lower lobe infiltrate (4) | Patchy and linear opacities in the bilateral mid and lower lung fields (7) | Bilateral mid and lower lung consolidations (12) | Bilateral patchy lower lung opacities (10) | Opacification of the left lower lung (7) |
| T<sub>max</sub>, °F (day) | 102.9 (7, 9, 11) | 100.8 (10) | 99.2 (4, 5) | 102.7 (9) | 99.1 (9) | 102.6 (10) | 102.4 (9) |
| Lowest SpO<sub>2</sub>, % (day) | 90 (10, 12) | 93 (23, 24) | 88 (4) | 87 (12) | 93 (13, 14) | 86 (12) | 92 (11) |
| Maximum O<sub>2</sub> support received, L/min | 2 | None | 4 | 20, HFNC | None | 3 | None |
| Peak AST, U/L (day) | 129 (13) | 46 (19) | 47 (11) | 99 (7) | 190 (13) | 167 (12) | 163 (14) |
| **Peak ALT, U/L (day)** | 219 (13) | 66 (23) | 75 (12) | 136 (6) | 389 (15) | 248 (14) | 127 (14) |
| **Lowest WBC count, cells/μL (day)** | 3300 (9) | 3000 (8, 12, 19) | 3400 (4) | 3900 (5) | 3900 (13) | 5700 (15) | 2400 (10) |
| **Highest procalcitonin, ng/mL (day)** | <0.05 (9, 11) | <0.10 (8) | Not tested | 0.21 (10) | 0.13 (13) | 0.13 (10, 11) | 0.07 (14) |
| **Molecular test results for other viral pathogens (day)** | PCR negative for Adenovirus; Coronavirus 229E/HKU1/NL63/OC43; HMPV; Influenza A & B; Parainfluenza 1–4; RSV; Rhino/enterovirus; (4) | PCR negative for Adenovirus; Coronavirus 229E/HKU1/NL63/OC43; HMPV; Influenza A & B; Parainfluenza 1–4; RSV; Rhino/enterovirus; (7) | PCR negative for Adenovirus; Coronavirus 229E/HKU1/NL63/OC43; HMPV; Influenza A & B; Parainfluenza 1–4; RSV; Rhino/enterovirus; (4) | PCR negative for Adenovirus; Coronavirus 229E/HKU1/NL63/OC43; HMPV; Influenza A & B; Parainfluenza 1–4; RSV; Rhino/enterovirus; (3) | PCR negative for Adenovirus; Coronavirus 229E/HKU1/NL63/OC43; HMPV; Influenza A & B; Parainfluenza 1–4; RSV; Rhino/enterovirus; (9) | PCR negative for Adenovirus; HMPV; Influenza A & B; RSV; Parainfluenza 1–3; Rhinovirus; (10) | PCR negative for Adenovirus; HMPV; Influenza A & B; RSV; Parainfluenza 1–3; Rhinovirus (7) |
| **Blood cultures (collection day)** | Negative (7, 8, 9) | Negative (7) | Negative (4) | Negative (10) | Not collected | Negative (10) | Negative (7) |
| **Antibiotics (days)** | Vancomycin (10–11)<br>Cefepime (10–12) | Levofloxacin (8–14) | Ceftriaxone (4–11)<br>Azithromycin (4–8) | Metronidazole (33–42) | None | Ceftriaxone (9)<br>Azithromycin (9) | None |
| **Indication for antibiotics** | Hospital-acquired pneumonia | Community-acquired pneumonia | Community-acquired pneumonia | *Giardia lamblia*<br>*Clostridioides difficile* | n/a | Community-acquired pneumonia | n/a |
| **Received remdesivir\*\*\* (days)** | Yes (11–15) | No | Yes (7–10) | Yes (11–20) | No | No | No |
| **Symptoms and signs following remdesivir** | Nausea; gastroparesis; elevated aminotransferase levels | n/a | Mild nausea and abdominal discomfort at start of infusion; elevated aminotransferase levels | Loose stools; one episode of bloody stool; elevated aminotransferase levels | n/a | n/a | n/a |
| **Other medications received (day(s); indication)** | Benzonatate capsules<br>Acetaminophen<br>Ibuprofen<br>Dextromethorphan<br>Guaifenesin<br>Ondansetron<br>Lorazepam | Methylprednisolone 40 mg IV Q8H (day 8–9)<br>Prednisone 40 mg PO daily (day 9–10)<br>Nystatin (day 15–24; oral thrush) | Methylprednisolone (40 mg IV x 1)<br>Prednisone 40 mg (PO x 1) (day 5–6; COPD exacerbation)<br>Furosemide (day 5–13);<br>Nystatin (day 10–13; oral thrush) | Oseltamivir (day 3–5; empiric treatment while COVID-19 test pending);<br>Furosemide (day 10–11; worsening hypoxia) | Acetaminophen PRN<br>Ibuprofen PRN<br>Melatonin PRN | Acetaminophen PRN | Acetaminophen PRN |

Abbreviations: n/a, not applicable; CXR, chest radiograph; $T_{max}$ , maximum daily temperature; $SpO_2$ , minimum daily oxygen saturation; HFNC, high-flow nasal cannula; AST, aspartate aminotransferase; ALT, alanine aminotransferase; WBC, white blood cell count; PRN, as needed
\* All reported days are illness day.
\*\* Patient 9 reported diarrhea one day before development of fever and cough.
\*\*\* Chest computed tomography scan; findings for other patients are reported from chest radiographs.
\*\*\*\* Remdesivir dose: 200mg IV once on day 1, then 100mg IV daily

**Figure 1:**
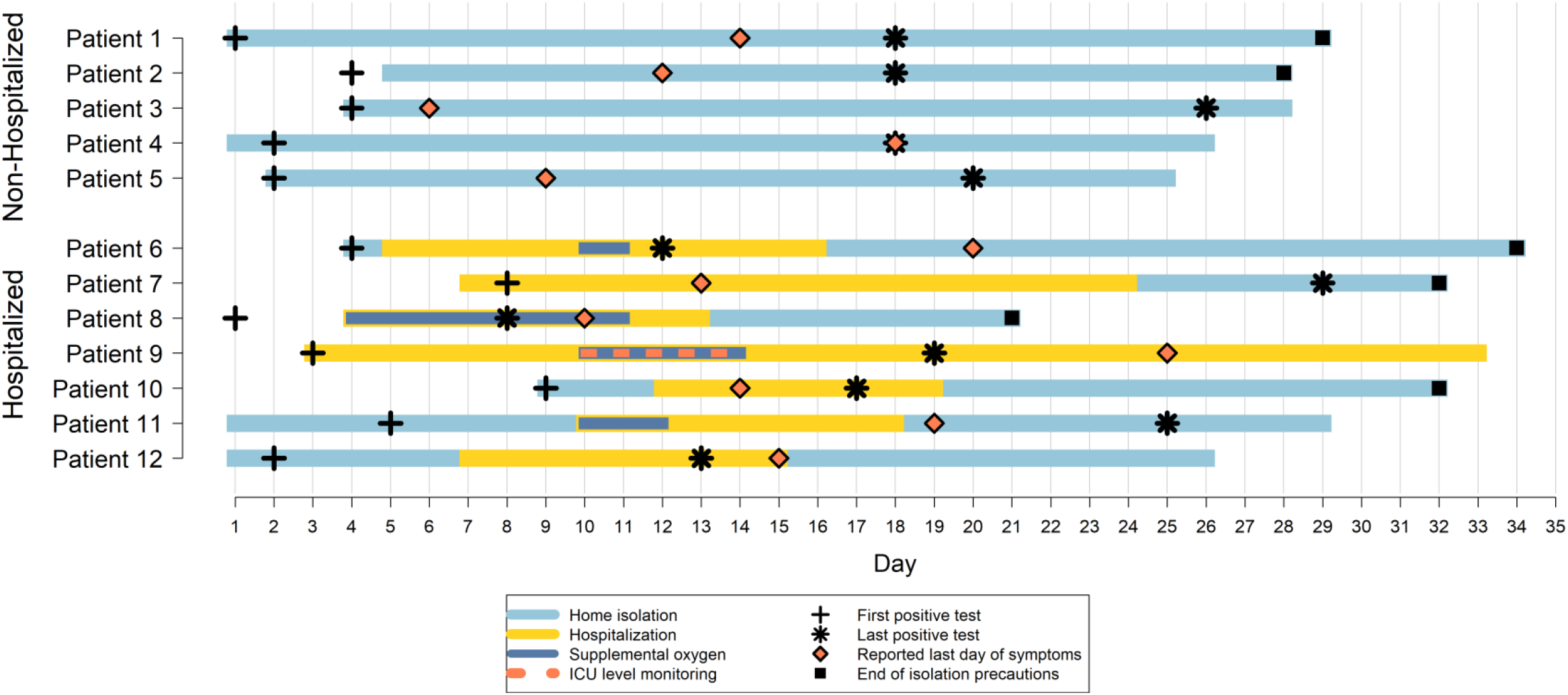
Timeline of illness onset, SARS-CoV-2 RNA detection, hospitalization, oxygen requirement, and reported symptom resolution among the first 12 patients with COVID-19 in the United States, January–February 2020. Patients 1 – 5 were not hospitalized and Patients 6 – 12 were hospitalized. Days are sequential from day of symptom onset (Day 1). Light blue bars indicate time patients were under home isolation. Yellow bars indicate duration of hospitalization. Dark blue bars indicate duration of supplemental oxygen administration in hospital. The orange dashed bar indicates duration of intensive care-level monitoring for Patient 9. The black “+” indicates collection date of the earliest sample that tested positive for SARS-CoV-2 by rRT-PCR. The black asterisk indicates collection date of the latest sample to test positive for SARS-CoV-2 by rRT-PCR. The orange diamond indicates date of last report of COVID-19-related symptoms. The black square indicates the last day of isolation precautions; patients with no black square were still under isolation precautions as of February 22. The last date of specimen collection was February 21, and the last date of testing was February 22. Patient 1 reported a cough with initial onset in mid-December before the patient traveled to China. The patient reported no change in the cough from the initial onset until reported resolution on day 18. Because onset date was difficult to determine for this patient, we have used date of detection as Day 1 to assess viral RNA detection.

Dates of illness onset ranged from January 14 through 29. Ten patients traveled to mainland China in the two weeks before illness onset, including nine to Wuhan City. Two patients’ only reported exposure was close contact with a previously identified U.S. patient with COVID-19. Among all patients, the duration of potential exposure ranged from 5 days to over 1 month; the time between last date of possible exposure and illness onset ranged from 0–5 days.

The most commonly reported signs or symptoms at illness onset were cough (n=8) and subjective or measured fever (n=7) (Table 1). Two patients reported neither fever nor cough at onset, though they did develop them subsequently: one reported diarrhea as the initial symptom (one day before fever and cough), and the other reported sore throat.

Over the course of illness, patients reported cough (n=12), subjective or measured fever (n=9), diarrhea (n=3), and vomiting (n=2). Three patients who did not report fever were never hospitalized and remained on home isolation. Of these, one patient reported only cough and rhinorrhea; one patient reported only cough which began before travel to China and did not change from the initial onset until resolution; and one patient reported cough, chills, fatigue, headache, and nausea.

### Clinical course of hospitalized patients

The clinical course for each hospitalized patient is described in the Appendix. The median duration of fever was 9 days (range: 2–11). Peak body temperature during hospitalization occurred at a median of illness day 9 (range: 4–10) (Figure 2). All hospitalized patients had oxygen saturation <94% on room air at some point during their illness, with the oxygen saturation nadir (range: 86–93%) occurring at a median of illness day 12 (range: 4–23) (Figure 2). Five patients reported difficulty breathing, and four received supplemental oxygen (Table 2; Figure 1). Patient 9 required high-flow nasal cannula oxygen supplementation and intensive care monitoring.

**Figure 2:**
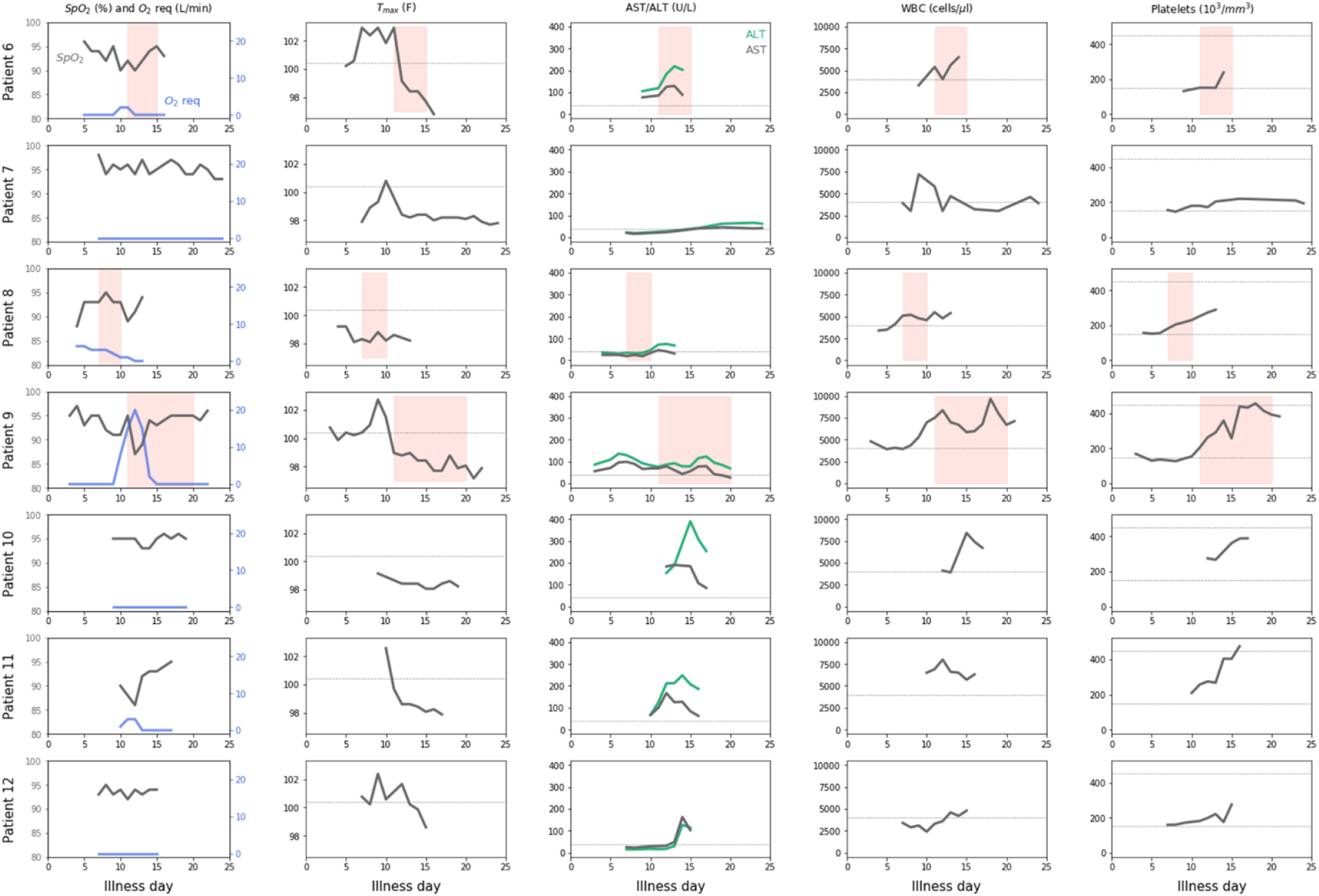
Clinical and laboratory values by illness day for the first seven patients hospitalized with COVID-19 in the United States, January–February 2020. Abbreviations: SpO_2_, oxygen saturation; O_2_ req, supplemental oxygen requirement; T_max_, maximum body temperature; AST, aspartate aminotransferase; ALT, alanine aminotransferase; WBC, white blood cell count. Pink shading: remdesivir administration. Dotted lines: 100.4 F (T_max_), 40 U/L (AST/ALT), 4000 cells/µl (WBC), 150 and 250 10^3^/mm^3^ (platelets)

Two patients received a short course (≤3 days) of corticosteroids. Three, including one who also received corticosteroids, received the investigational antiviral remdesivir (Gilead Sciences, Foster City, California) under expanded access (compassionate use) for a duration of 4–10 days. Following remdesivir initiation, all had transient gastrointestinal symptoms, including nausea, vomiting, gastroparesis, or rectal bleeding. No other post-remdesivir symptoms were observed. Patient 9 reported loose stool and rectal bleeding and had traveled in Mexico before illness onset; stool later tested positive for *Giardia* and *Clostridiodes difficile*. Remdesivir was discontinued after improvement in each patient’s respiratory symptoms.

Blood cultures were negative in 6/6 hospitalized patients tested, including those obtained from four patients treated empirically for bacterial pneumonia. Molecular testing for influenza A and B on respiratory specimens was negative, and multi-pathogen respiratory PCR panels were negative for all targets in all hospitalized patients (Table 2).

### Laboratory and radiographic findings among hospitalized patients

Six of seven hospitalized patients had leukopenia (<4000 cells/µl), and the white blood cell count nadir occurred at a median of illness day 9 (range: 4–15) (Figure 1). Procalcitonin levels were <0.15 ng/ml in five of six patients who had levels checked. Aminotransferase levels were elevated in all hospitalized patients: AST levels peaked (median peak value 129 U/L, range 46–190 U/L) at a median of illness day 13 (range 7–19) and ALT levels peaked (median peak value 136 U/L, range 66–389 U/L) at a median of illness day 14 (range: 6–23). Three of seven hospitalized patients had mild elevations in alkaline phosphatase levels >115 U/L (maximum value 163 U/L). Elevated lactate dehydrogenase levels >600 U/L, coinciding with clinical deterioration, were observed in two patients tested. No major elevations in serum total bilirubin (7 patients tested) or prolongations in prothrombin time (4 patients tested) were identified. Among the three remdesivir recipients, aminotransferase elevation developed in Patient 6 one day after starting remdesivir and in Patient 8 four days after starting remdesivir. Patient 9 had aminotransferase elevation at illness days 6–7 before starting remdesivir; aminotransferase levels started to decrease but increased again five days after starting remdesivir.

Unilateral or bilateral pulmonary opacities were seen on chest imaging at some point for all seven hospitalized patients (Table 2). Four hospitalized patients did not have any abnormalities identified on initial chest radiograph (illness day range: 4–9). Patient 7 had an abnormal chest computed tomography scan on the day of the normal chest radiograph (day 7).

### Virologic testing

#### Initial SARS-CoV-2 testing

All 12 patients had respiratory specimens collected between illness days 1–9 (median, day 4), and all tested positive in ≥ 2 specimen types (Figure 3). Among initial diagnostic specimens, SARS-CoV-2 RNA was detected in OP (11/11 patients), NP (10/12 patients), and sputum (4/4 patients). Viable SARS-CoV-2 virus was cultured from 5/6 initial NP specimens, 4/7 initial OP specimens, 3/3 initial sputum specimens, and 1/1 additional sputum specimen collected 3 days after the initial specimens (Figure 3). Virus was cultured from two patients who were not hospitalized (Figure 3).

**Figure 3.**
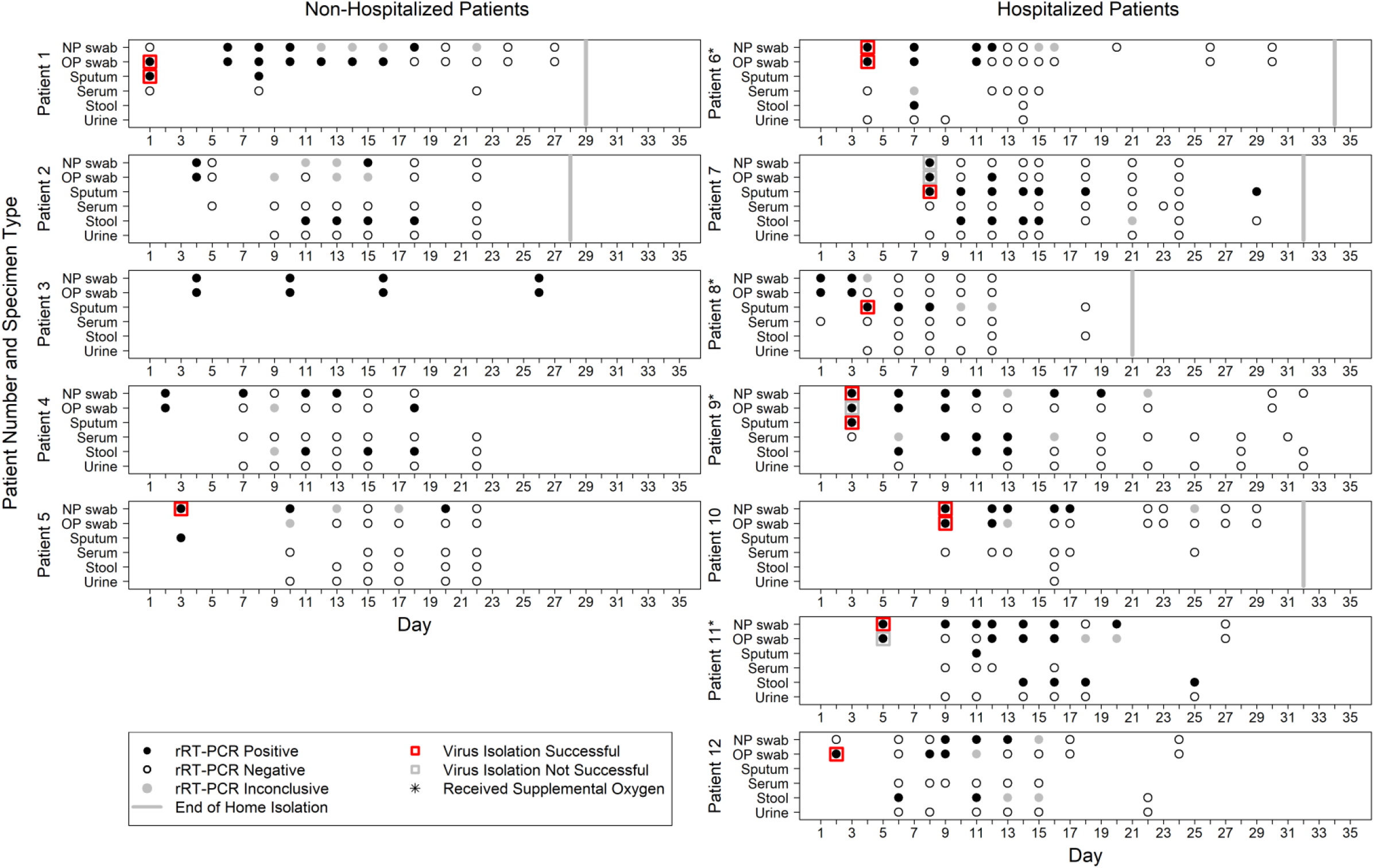
SARS-CoV-2 rRT-PCR results by specimen type and day among the first 12 patients with COVID-19 in the United States, January–February 2020. Specimen types tested include nasopharyngeal (NP) swab, oropharyngeal (OP) swab, sputum, serum, stool, and urine. Day of illness is the number of days from the date of symptom onset (day 1) until the date of specimen collection. Virus culture was attempted on selected respiratory specimens collected early in the course of illness. rRT-PCR results were reported as positive (all three targets positive), negative (all three targets negative), or inconclusive (only one or two positive targets). Red squares surrounding black-filled circles indicate rRT-PCR-positive specimens from which virus culture was successful. Gray squares surrounding black-filled circles indicate rRT-PCR-positive specimens from which virus culture was unsuccessful. Black-filled circles indicate rRT-PCR-positive specimens. Black-outlined circles indicate rRT-PCR-negative specimens. Gray-filled circles indicate specimens with inconclusive rRT-PCR results. The gray vertical bar indicates the date home isolation was discontinued. An asterisk indicates patients who required supplemental oxygen.

#### Serial SARS-CoV-2 testing

As of February 22, 398 specimens were collected and tested from the 12 patients throughout the course of illness. All 12 patients had SARS-CoV-2 RNA detected in at least one NP swab, 11/12 in an OP swab, 6/6 in sputum, 1/12 in serum, 7/10 in stool, and 0/10 in urine (Figure 3). Among 98 pairs of simultaneous NP and OP specimens, 58 (59%) had concordant results. Among 27 discordant pairs with one positive specimen, the NP specimen was positive in 70%; the remaining 13 discordant pairs had one negative and one inconclusive specimen. Two patients provided sputum specimens when NP and/or OP specimens tested negative, and sputum continued to be positive in both patients. In Patient 7, viral RNA was detected in sputum 17 days after the last positive OP specimen and ≥ 2 weeks after reported symptom resolution. In seven patients who had SARS-CoV-2 RNA detected in stool, most detections occurred when viral RNA was still detectable in the respiratory tract. Among three patients who reported diarrhea, all had viral RNA detected in stool.

Mean Ct values in positive specimens were 17.0–39.0 for NP, 22.1–39.7 for OP, and 24.1–39.4 for stool. Ct values were lower in the first week of illness than the second in most patients (Figure S1); in some patients, low Ct values continued into the 2^nd^ and 3^rd^ week of illness. There was no apparent relationship between Ct values in the upper respiratory tract and disease progression. SARS-CoV-2 rRT-PCR results turned positive in serum of Patient 9 in the second week of illness at the time of rapid clinical deterioration.

Serial testing to determine duration of RNA detection and viral shedding is ongoing. As of February 22, SARS-CoV-2 RNA has been detected at a maximum of day 26 in NP specimens, day 26 in OP, day 29 in sputum, and day 25 in stool (Figure 3). The duration of viral RNA detection did not differ by hospitalization status or supplemental oxygen requirement.

### Outcomes

As of February 22, all patients reported symptom resolution (Figure 1). Eleven patients reported cough, (often intermittent) as the last symptom. Median symptom duration was 14 days (range: 6–20). SARS-CoV-2 RNA was detected after reported symptom resolution in 7/11 patients, including in NP (n=6), OP (n=2), sputum (n=1), and stool (n=3) specimens. As of February 22, one patient remained hospitalized, and five patients remained on home isolation. Home isolation was discontinued for 6 patients per CDC criteria;^14^ the last respiratory specimens with a positive or inconclusive test result were collected from these patients on days 12–29.

### Genome sequencing and phylogenetic analysis

Complete genome sequences were generated from respiratory specimens from all 12 patients. The sequences had >99% nucleotide identity to 85 reference sequences of SARS-CoV-2 genomes; phylogenetic tree analysis identified a few distinct subgroups (Figure S2) which were not divergent from each other, indicating that the outbreak may still be in an early stage.

## DISCUSSION

We describe the first 12 patients with confirmed COVID-19 in the United States, including clinical course of the first 7 hospitalized patients. Nine patients had traveled to Wuhan City, the epicenter of the outbreak, one had traveled to China but not to Hubei Province, and two had close contact with a patient with confirmed COVID-19 in the United States, representing domestic human-to-human transmission. Illness ranged from mild to moderately severe, and hospitalized patients showed signs of clinical worsening in the second week. All patients recovered or are improving, and three patients tolerated treatment with the investigational antiviral remdesivir. SARS-CoV-2 RNA was detected in upper and lower respiratory specimens, stool, and serum. The highest viral RNA levels were detected in upper respiratory tract specimens, typically during the first week of illness. SARS-CoV-2 was cultured from the initial respiratory specimens of mild and moderately ill patients. Viral RNA was still detected after reported symptom resolution for seven patients. SARS-CoV-2 genome sequencing and phylogenetic analysis from these 12 patients’ respiratory tract specimens support a single recent zoonotic transmission event in Wuhan City and subsequent human-to-human transmission.

Overall, these patients had milder disease than those in initial reports from China describing higher rates of complications and death.^15–18^ Initial case identification in China focused on hospitalized patients with pneumonia, but recent reports have described a milder clinical course, consistent with our findings.^19–23^ Among hospitalized patients in this report, the second week of illness was characterized by clinical or laboratory signs of worsening such as hypoxemia, increase in fever, or elevation of aminotransferases. Although some patients received empiric antibiotic treatment for possible secondary bacterial pneumonia, no definitive evidence of bacterial co-infection was found. Worsening in the second week of illness is consistent with previous reports^15,17^ and highlights the importance of close monitoring beyond the first week of illness, even in patients with mild illness or no initial radiographic abnormalities.

Patient 9, the most severely ill among this series, experienced sudden clinical deterioration late in the second week of illness. This was the only patient with SARS-CoV-2 RNA detected in serum, and detection in serum was temporally related to clinical deterioration. Similar observations have been described previously.^24,25^ Increased proinflammatory cytokines have been observed in patients with COVID-19,^17^ and it is possible that cytokine dysregulation and endothelial dysfunction contribute to both clinical worsening and SARS-CoV-2 RNA detection in serum.

Characterizing SARS-CoV-2 shedding is important to understand transmission and guide prevention strategies. We detected viral RNA and cultured virus from upper respiratory specimens, even from patients with lower respiratory tract illness. In general, Ct values in upper respiratory tract specimens were lowest during the first week of illness (suggesting high RNA counts), consistent with previous reports.^25–27^ SARS-CoV-2 RNA was detected in respiratory tract specimens for 2–3 weeks in most patients and for up to 29 days as of February 22. In two patients with a productive cough, viral RNA was detected in sputum after RNA was no longer detectable in NP or OP specimens. SARS-CoV-2 RNA levels and duration of RNA detection did not appear to vary by illness severity, and several patients had viral RNA detected in respiratory specimens after reported symptom resolution.

We detected SARS-CoV-2 RNA in stool of multiple patients; testing continues to assess if this represents viable virus. SARS-CoV-2 was cultured from one patient’s stool in China^28^ but the implications for transmission are unclear. We detected SARS-CoV-2 RNA in the serum of one hospitalized patient but did not detect RNA in urine. More data are needed to better understand how duration of RNA detection, RNA levels, and viable virus are related to symptom progression, illness severity, and transmission.

Three hospitalized patients received the investigational antiviral remdesivir under expanded access (compassionate use) at the time of clinical worsening based upon a decision by each patient’s clinician. Remdesivir inhibits viral replication through premature termination of RNA transcription.^29,30^ *In vitro* studies have demonstrated that remdesivir inhibits SARS-CoV-2 replication in non-human cells.^31^ Because remdesivir use was not given as part of a randomized controlled trial, we are unable to assess its effectiveness or safety. Randomized controlled trials of remdesivir are underway.^32–34^ Two hospitalized patients received corticosteroids. WHO interim guidance for clinical management of severe acute respiratory infection with suspected COVID-19 advises against use of corticosteroids unless indicated for another reason.^35^

Several limitations should be considered when interpreting our findings. Our sample of patients is small. Information collected from patient interviews may have been subject to response bias. The threshold for admitting patients and for monitoring in the hospital was likely lower than for other respiratory infections because of uncertainty about the clinical course of COVID-19. Dates of illness resolution may be imprecise due to non-specific lingering symptoms or symptoms from chronic or unrelated conditions. Clinical laboratory tests and radiographic studies were ordered as a part of routine patient care and were not collected systematically. SARS-CoV-2 RNA detection does not necessarily reflect the presence of infectious virus, and rRT-PCR Ct values may have varied due to specimen collection or handling. Specimen collection is ongoing to inform both clinical management and infection prevention and control practices, and findings will be updated as more information becomes available.

## Conclusions

Characterization of the first 12 patients with COVID-19 identified in the United States, including 7 hospitalized patients, provides key insight into the epidemiology, clinical characteristics, and natural history of SARS-CoV-2 infection. These patients experienced mild to moderately severe illness. Clinicians should anticipate that some patients may worsen in the second week of illness. Early and prolonged detection of SARS-CoV-2 RNA suggest the window for diagnosis of COVID-19 is long. Although duration of infectiousness is unclear, our early data show viable virus can be cultured readily from upper respiratory tract specimens soon after illness onset. Further investigations are needed to understand clinical course, immunologic response, SARS-CoV-2 RNA detection, virus culture, and transmission, to inform clinical management and public health strategies to prevent disease spread.

## Data Availability

Data are available upon request.

## Disclaimer

The findings and conclusions in this report are those of the author(s) and do not necessarily represent the official position of the Centers for Disease Control and Prevention.

